# Cardiac or Infectious? Transfer Learning with Chest X-Rays for ER Patient Classification

**DOI:** 10.1101/2020.04.11.20062091

**Authors:** Jonathan Stubblefield, Mitchell Hervert, Jason Causey, Jake Qualls, Wei Dong, Lingrui Cai, Jennifer Fowler, Emily Bellis, Karl Walker, Jason H. Moore, Sara Nehring, Xiuzhen Huang

**Affiliations:** The Joint Translational Research Lab of Arkansas State University and St. Bernards Medical Center, Jonesboro, Arkansas, 72467; The Internal Medicine Residency Program, St. Bernards Medical Center, 225 E Jackson Ave, Jonesboro, Arkansas, 72401; Institute for Biomedical Informatics, University of Pennsylvania, Philadelphia, Pennsylvania, 19104; Ann Arbor Algorithms, Ann Arbor, Michigan 48103, United States of America; Department of Mathematics and Computer Science, University of Arkansas at Pine Bluff, Pine Bluff, Arkansas 55455; Department of Computer Science and Molecular Biosciences Program, Arkansas State University, Jonesboro, Arkansas 72467; Arkansas Biosciences Institute, Arkansas State University, Jonesboro, Arkansas 72467

**Keywords:** Emergency room (ER) patient classification, Chest x-ray, Deep-learning model

## Abstract

One of the challenges with urgent evaluation of patients with acute respiratory distress syndrome (ARDS) in the emergency room (ER) is distinguishing between cardiac vs infectious etiologies for their pulmonary findings. We evaluated ER patient classification for cardiac and infection causes with clinical data and chest X-ray image data. We show that a deep-learning model trained with an external image data set can be used to extract image features and improve the classification accuracy of a data set that does not contain enough image data to train a deep-learning model. We also conducted clinical feature importance analysis and identified the most important clinical features for ER patient classification. This model can be upgraded to include a SARS-CoV-2 specific classification with COVID-19 patients data. The current model is publicly available with an interface at the web link: http://nbttranslationalresearch.org/.

**Data statement:** The clinical data and chest x-ray image data for this study were collected and prepared by the residents and researchers of the Joint Translational Research Lab of Arkansas State University (A-State) and St. Bernards Medical Center (SBMC) Internal Medicine Residency Program. As data collection is on-going for the project stage-II of clinical testing, raw data is not currently available for data sharing to the public.

**Ethics:** This study was approved by the St. Bernards Medical Center’s Institutional Review Board (IRB).

## 1. Background

In this study, we focused on acute respiratory distress syndrome (ARDS) in an emergency room (ER) setting. There are many etiologies of acute dyspnea, but our model focused on distinguishing between two major categories: cardiac and infectious. Upon admission to a hospital emergency department, attending physicians must quickly determine which category the patient falls into. Typically these patients receive a suite of common clinical panels as well as a chest X-ray image early in the diagnostic process. We have developed a machine learning model capable of assisting ER physicians with categorizing the acute dyspnea given clinical values alone, or in conjunction with an X-ray image if one is available.

Cardiac causes include etiologies of dyspnea secondary to a misfunction in the heart, including acute coronary syndrome, acute heart failure, arrythmias, and valvular disease [1]. These diseases do not benefit from antibiotic therapy. Infectious causes of acute dyspnea include both pneumonia, an infectious process primary to the lungs, and sepsis, a systemic response to an infection anywhere in the body that can impair function in a variety of organs [1]. When severity is life-threatening, suspicion of these disease processes require empiric antibiotics. Other causes of acute dyspnea may fall into neither of these categories [1], or a patient may have acute dyspnea with both cardiac and infectious contributions. See Figure 1 for examples of X-ray images for patients with neither label, “infection” label, and “cardiac” label, respectively.

**Figure 1:**
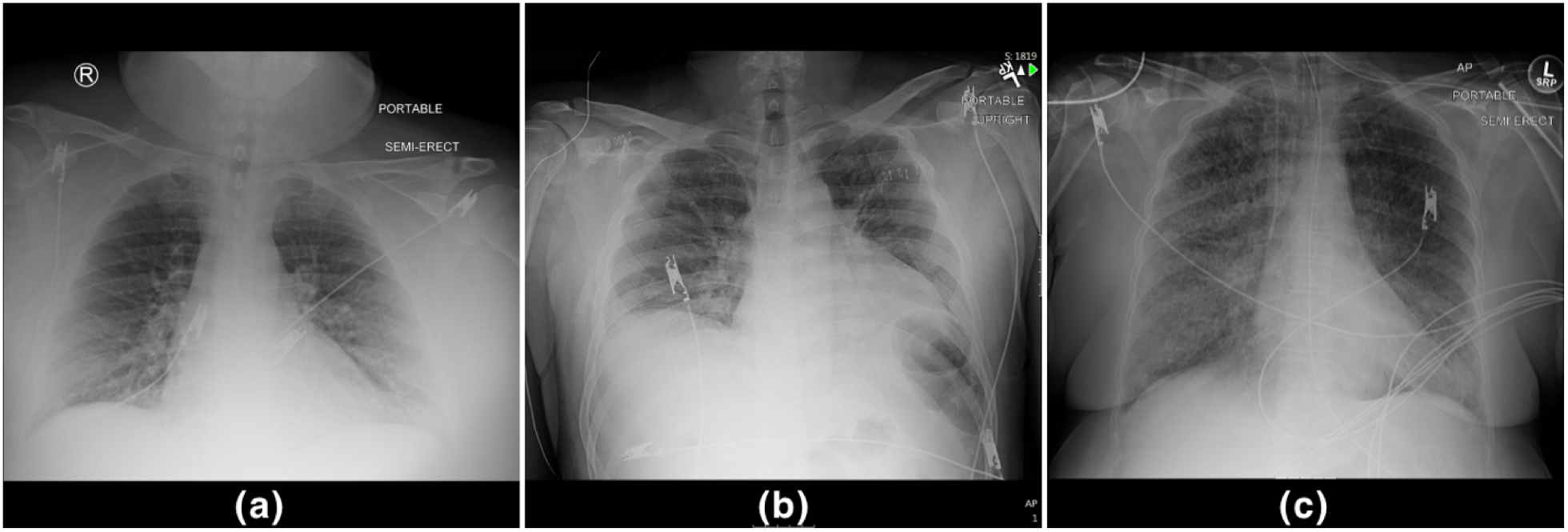
Examples of chest X-ray images for (a): neither infection nor cardiac label, (b): infection label, (c) cardiac label.

Antibiotics are not benign medications. Though they are safe and efficacious when used judiciously, incautious use of antibiotic causes many problems for the patient and society as whole. Antibiotics have adverse effects in 1 out 5 patients [9]. Though these adverse effects often include direct side-effects from the drugs themselves, antibiotics can also produce adverse effects through their interactions with microorganisms [9]. Misuse of antibiotics can promote the growth of antibiotic resistant populations or cause an overgrowth of an opportunistic pathogen, such as *Clostridium dificile* [9]. However, in an ER setting, empiric antibiotics may need to be started without confirmation of infectious process that would benefit from antibiotic therapy [9]. In these instances, the risk of harm from not initiating antibiotics promptly is greater than the risk posed by the antibiotics themselves. This is largely due to the difficulty in excluding infectious processes from the diagnosis. Excluding serious infectious etiology of disease usually requires taking samples from the patient and waiting several days to see if bacteria grow from the sample. Our model represents a step toward more rapidly confirming or excluding infectious etiology, and thus reducing the unnecessary empiric prescription of antibiotics.

Recent advances in machine learning have shown that it can be a valuable tool for aiding in medical planning. The CheXNet [14] model was able to accurately identify 14 categories of abnormalities in chest X-ray images. Deep learning techniques have shown promise for automated detection and diagnosis of lung cancer [19-22], breast cancer [23,24], skin cancer [25-27], and other diseases. Most of these approaches use deep neural networks [28] especially convolutional neural networks [29,30]. Gradient boosted trees, and the XGBoost [3] model in particular, have been used to solve a wide variety of problems where the inputs may include very diverse variables of differing types [31-34]. Our model makes use of both deep neural networks and XGBoost for examining images and clinical data, respectively, and the combination of the two is handled by extracting image features via a deep neural network and performing classification using XGBoost.

## 2. Methods

### 2.1 Clinical Data Preprocessing

The dataset contains clinical data of 188 patients and chest X-ray images of 171 patients. Each patient has two boolean classification labels: cardiac and infection, of which both can be true. We used the clinical and image data of the 171 patients who had both for evaluation. The clinical data were hand-entered by a group of residents on rotation and contained some data entry errors that required careful cleaning before it could be used.

The complete blood count (CBC) with differential column always contained 3 or 4 values. Based on what CBC with diff reports, conventional notation, and their ranges, these values were white blood cell count, hemoglobin, hematocrit, and platelets, respectfully. When three values were present, hematocrit was always assumed to be missing based again on ranges and conventions. Of note, hematocrit should be able to be calculated from hemoglobin and is somewhat of a “redundant” value. After cleaning, hematocrit was excluded from the final analysis due to a preponderance of missing values.

The basic metabolic profile (BMP) test reports sodium, potassium, chloride, bicarbonate, blood urea nitrogen, and glucose. By convention, they are reported in this order. The original dataset included some missing values in the BMP report. We identified which values were missing based on the positions and ranges of the values present compared to typical ranges for corresponding components of the BMP.

The column for brain natriuretic peptide (B-NP) always contained a single value. Where a real number was present, the value was kept as is. Otherwise, it was given an appropriate sentinel value to represent “missing”.

The first troponin measurement was represented as a continuous (real number) value, but sometimes contained values that could be directly interpreted as a real number. Values such as “<0.012” were given the sentinel value “0” for “undetectable.” Multiple values were sometimes given, documenting the trend of multiple troponin measurements. In these cases, only the first measurement was kept.

The procalcitonin measurements contained too many missing values to be used in the final analysis, so it was excluded.

The lactic acid value was measured as a continuous (real number) value. All instances containing a value that could be directly interpreted as a real number were kept. All other values were marked as “missing”.

The vital signs column usually contained 6 values in the following format: Temperature; Pulse Rate; Systolic Blood Pressure/Diastolic Blood Pressure; Respiration Rate; Pulse Oximetry. Real number values were recorded without lettering or comments. Though pulse oximetry is typically recorded as a percentage, we converted it to a real number in the range [0,1]. The residents recording these measurements were not consistent with the ordering of these values. The overwhelming common alternative format transposed the blood pressure and pulse rate values. Missing values were identified using typical ranges of these values, the order of the values, and the fact that blood pressure values are always expressed as x/y with x>y. Information about the patient’s use of supplemental oxygen was not kept.

The arterial blood gas column was the most problematic. There were usually 4 or 5 values: Arterial pH, arterial pressure of CO2 (PCO2), arterial bicarbonate (bicarb), arterial pressure of O2 (PO2), and pulse oxygenation (SpO2) at time of blood draw. The residents recording these measurements were least consistent following the conventional order for this column. The conventional order of pH PCO2, bicarb, PO2, SpO2 was assumed unless the values were outside the typical range. However, interpretation was limited as the typical and possible ranges of PCO2 and PO2 overlap significantly. Though recommended for proper interpretation, information on SpO2 and patient supplemental oxygen utilization were not included.

### 2.2 CheXNet-Based Image Features

Training a deep convolutional neural network model requires a large number of images. In this dataset we have 171 images, which is too few to attempt training a complex and robust model from scratch. Instead, we opted to use a pre-trained neural network model from a similar application area as a feature extractor. CheXNet [14] is a 121-layer convolutional neural network trained on the NIH ChestX-ray14 [17] dataset, consisting of 100,000 frontal X-ray images with 14 disease labels. We used the open source PyTorch implementation of CheXNet [18].

We utilize the 14 output scores produced by the output stage of the pre-trained CheXNet model as 14 image features, and performed testing to determine whether adding these image features to the clinical features could improve classification accuracy. The CheXNet output scores are real number values in the range [0,1] and were originally interpreted as the probability that the input chest X-ray image should be labeled with the corresponding medical condition. We interpret the values as a 14-dimensional feature vector which is concatenated to our clinical features. The rationale is that this feature vector contains a high-level encoding of the medically relevant abnormalities observed in the X-ray image.

### 2.4 Model Training and Evaluation

We used XGBoost [3], an open-source implementation of gradient boosted decision trees. The model was trained and evaluated on the dataset using 5-fold cross-validation.

### 2.5 SHAP Analysis of Feature Importance

SHAP (SHapley Additive exPlanations) is a game theoretic approach to machine learning model explanation [10]. We used the Python implementation [11].

The Shapley analysis computes the Shapley value to each individual feature of a training sample. The Shapley value represents a feature’s responsibility for a change in the model’s output. We therefore use the sum of the magnitudes of the SHAP values across training examples to measure the importance of a feature.

## 3. Results

### 3.1 Separate Clinical, Image Models vs Clinical + Image Model

#### 3.1.1 Infection

Table 1 shows the performance on the “infection” labeling task for each fold of the 5-fold cross validation, as well as the average for all five folds, for three variations of the models. The “clinical” column corresponds to a model using clinical features only, which achieved an average accuracy of 63.8%. The “image” column corresponds to a model using image (CheXNet) features only, with an average accuracy of 63.8%. The “both” column corresponds to a model combining the clinical and image features, and had an average accuracy of 67.5%, which was a slight (3.7%) improvement over either of the single-modality models alone. Figure 2 shows a boxplot representing the range of accuracy values over the five folds.

**Table 1:** Five-fold cross validation results for “infection” label, showing clinical-only, image-only, and combined performance. Each fold is shown, with average performance in the last row. Top performance in each row is shown in bold.

| fold | clinical | image | both |
| --- | --- | --- | --- |
| 1 | 0.542 | **0.625** | 0.621 |
| 2 | 0.629 | 0.525 | **0.632** |
| 3 | 0.731 | 0.787 | **0.850** |
| 4 | **0.688** | 0.628 | 0.628 |
| 5 | 0.600 | 0.623 | **0.642** |
| **avg** | 0.638 | 0.638 | **0.675** |

**Figure 2:**
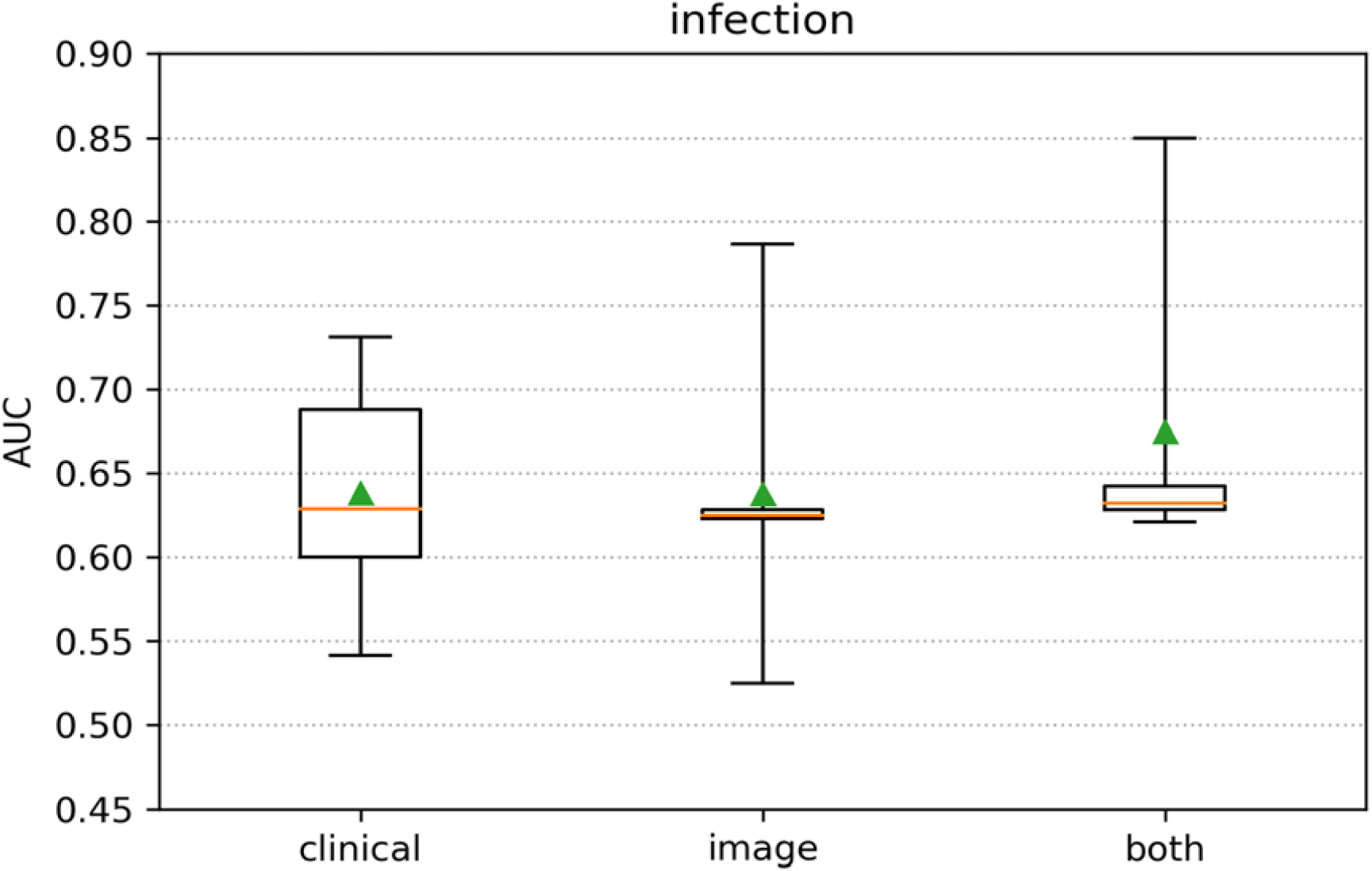
Box plot of 5-fold cross validation results. The box extends from the lower to upper quartile values of the data, with a line at the median and a triangle at the mean.

#### 3.1.2 Cardiac

Table 2 shows the performance on the “cardiac” labeling task for each fold of the 5-fold cross validation, as well as the average for all five folds, for three variations of the models. The “clinical” column corresponds to a model using clinical features only, which achieved an average accuracy of 70.3%. The “image” column corresponds to a model using image (CheXNet) features only, with an average accuracy of 59.7%. The “both” column corresponds to a model combining the clinical and image features, and had an average accuracy of 74.6%, which was a 4.3% improvement over either of the single-modality models alone. Figure 3 shows a boxplot representing the range of accuracy values over the five folds.

**Table 2:** Five-fold cross validation results for “cardiac” label, showing clinical-only, image-only, and combined performance. Each fold is shown, with average performance in the last row. Top performance in each row is shown in bold.

| fold | clinical | image | both |
| --- | --- | --- | --- |
| 1 | 0.720 | 0.747 | **0.793** |
| 2 | 0.677 | 0.535 | **0.719** |
| 3 | 0.618 | 0.628 | **0.715** |
| 4 | 0.743 | 0.528 | **0.795** |
| 5 | **0.754** | 0.547 | 0.705 |
| avg | 0.703 | 0.597 | **0.746** |

**Figure 3:**
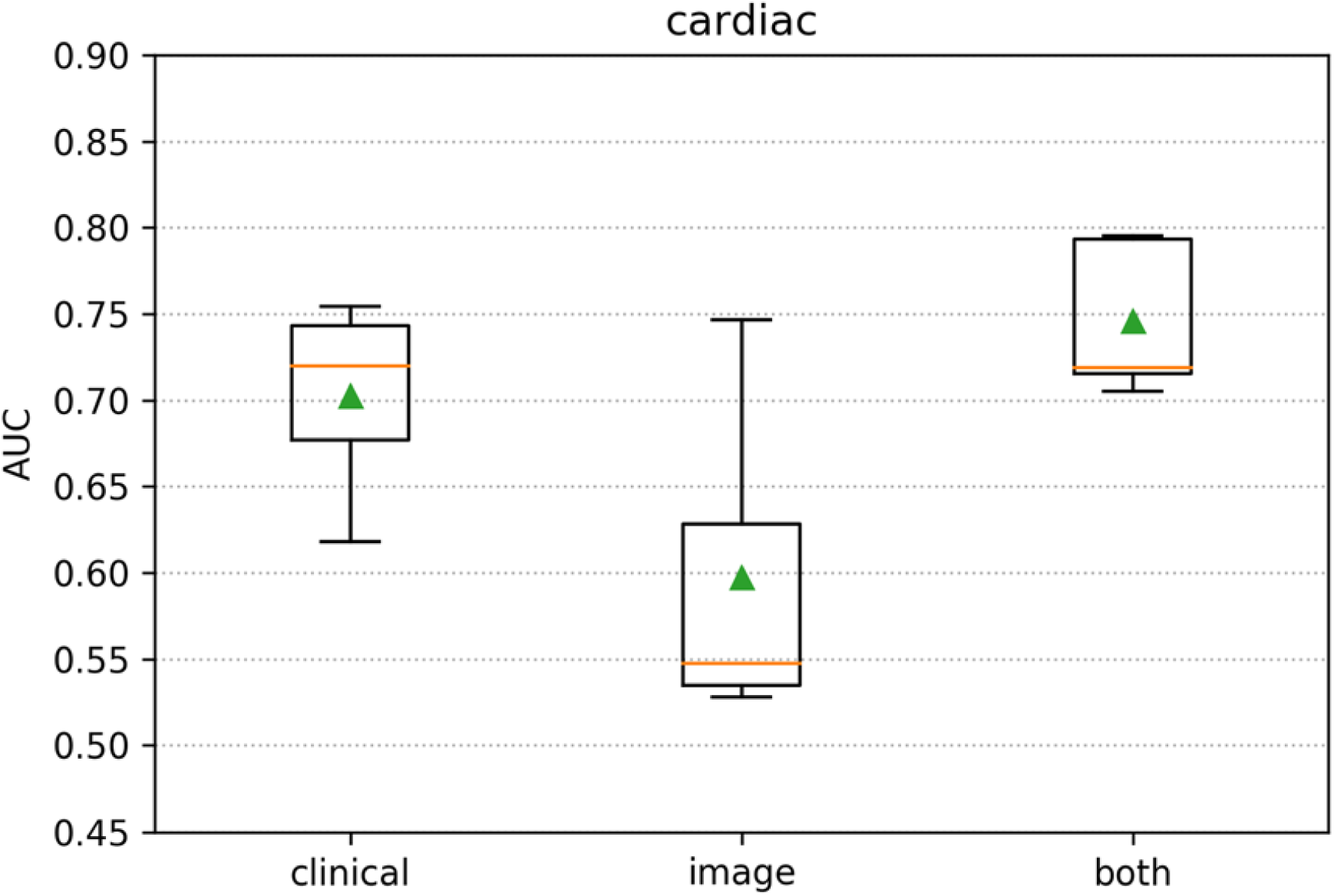
5-fold cross validation scores. The box extends from the lower to upper quartile values of the data, with a line at the median and a triangle at the mean.

### 3.2 Feature Importance Analysis

#### 3.2.1 Infection

Figure 4 shows the SHAP feature importance analysis for clinical features on the “infection” labeling task, and Figure 5 shows the same analysis for image features listed by categorical label as defined by CheXNet [14]. Each point in the figure is a feature value of a particular training example. All 171 examples are used for the Shapley analysis. The color of the point represent the feature value and the X axis of the point is its SHAP value. The features are ranked by the sum of SHAP value magnitudes over all samples.

**Figure 4:**
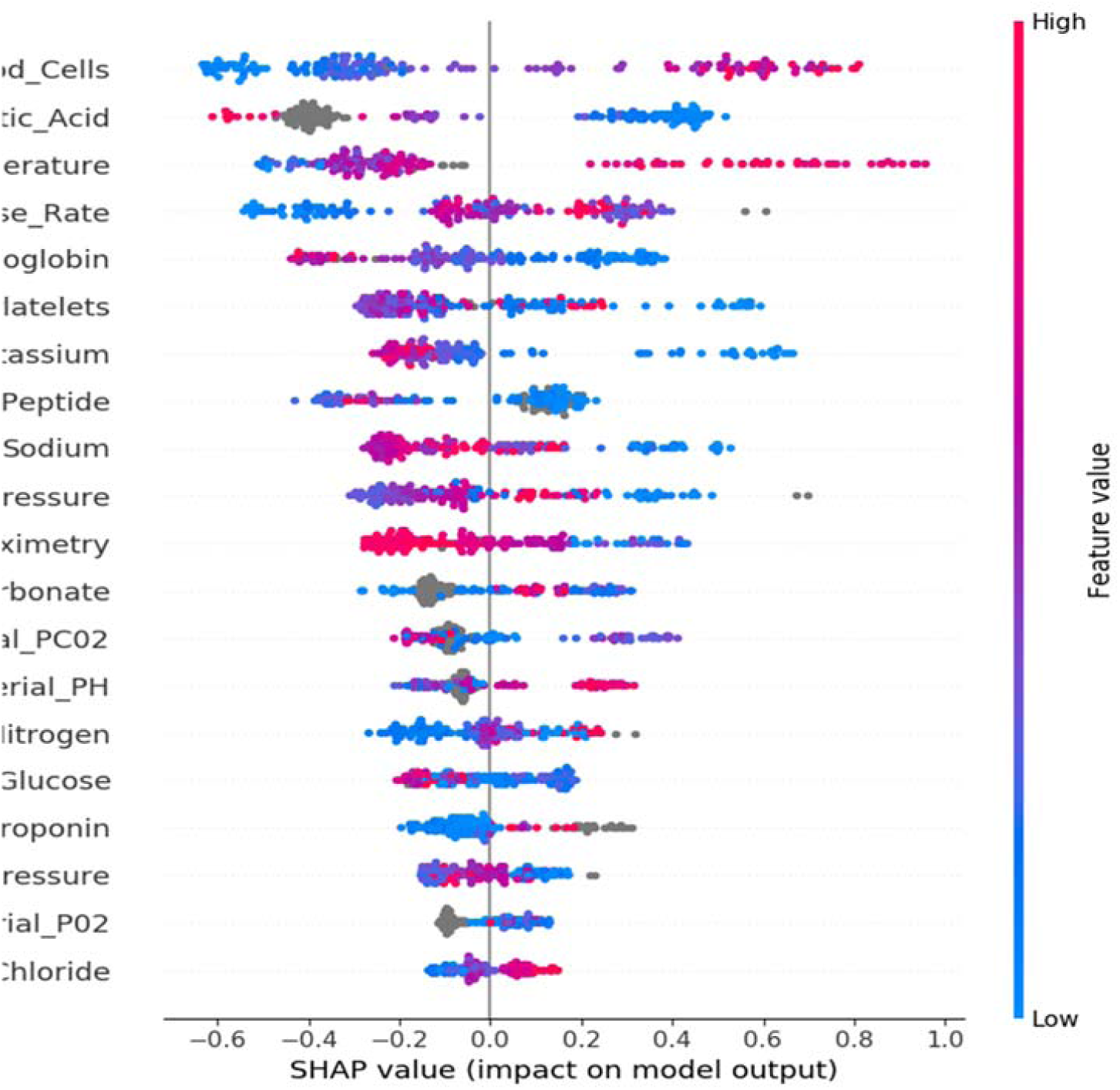
SHAP Feature importance plot for clinical features on the “infection” labeling task.

**Figure 5:**
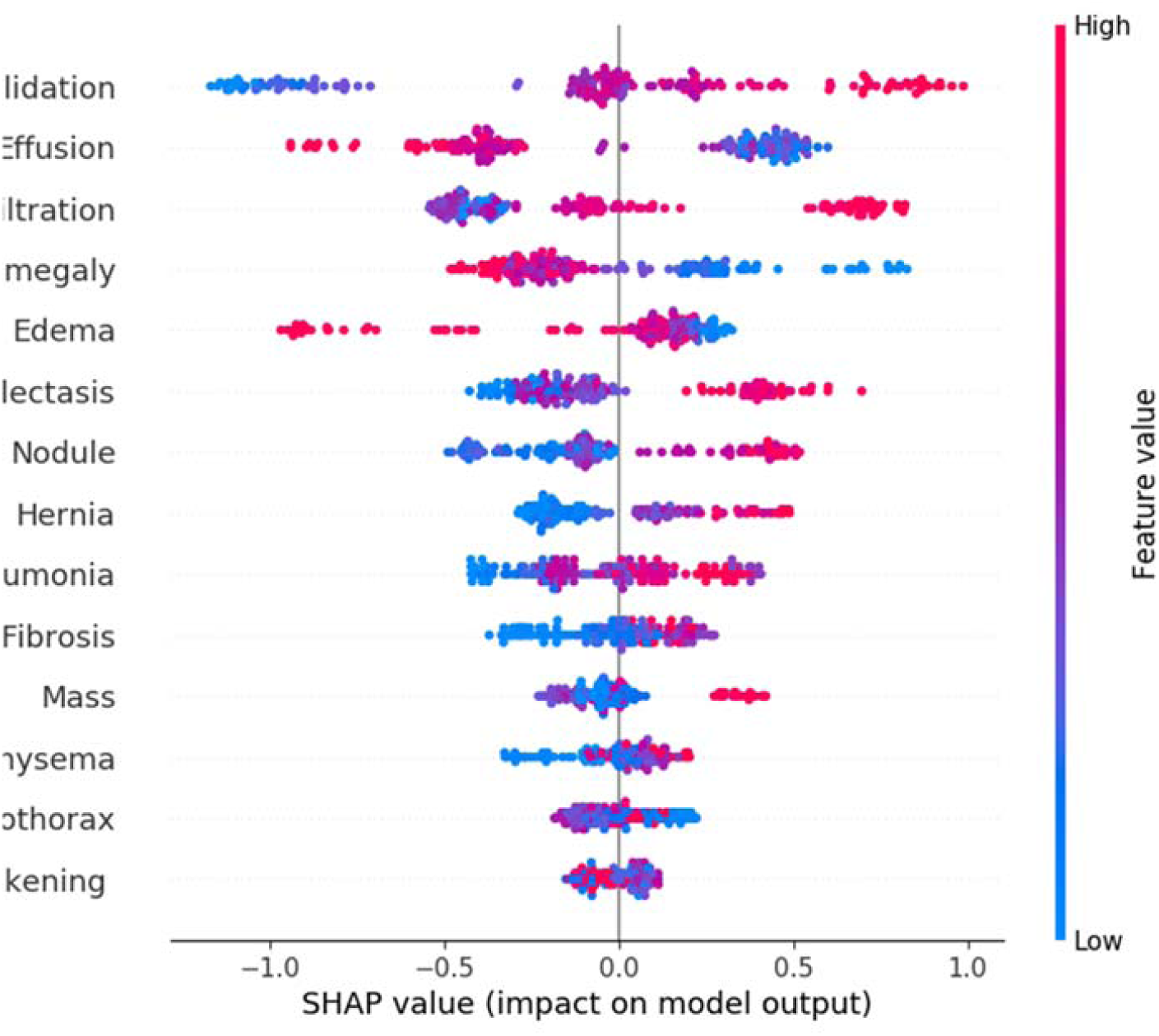
SHAP Feature importance plot for image features on the “infection” labeling task.

SHAP analysis of feature importance for prediction of infection was consistent with current medical knowledge. White blood cells count is expected to rise in response to infection [4] and was found to be the most important feature for determining the presence of infection. Similarly, fever (elevation of temperature) was found to be predictive of infection.

SHAP analysis of imaging features for prediction of infection is also consistent with clinical knowledge. Consolidation and infiltration can both be radiographic features of a pneumonia [2].

#### 3.2.2 Cardiac

SHAP analysis of feature importance for prediction of a cardiac cause of respiratory distress also followed a reasonable pattern (see Figures 6 and 7). Brain natriuretic peptide (B-NP) was rated by the model as its most important feature for predicting a cardiac cause of infection. Normal values for B-NP have been shown to have a high negative-predictive value for heart failure [5] and are used to diagnose exacerbation of existing heart failure [13]. Blood glucose levels are not directly associated with heart failure, but the model may be looking for associated diabetes mellitus. This common disease is an important risk factor for heart disease [6]. Increases in respiratory rate can be caused by heart failure exacerbations [13], so it makes sense that this would be an important predictive feature.

**Figure 6:**
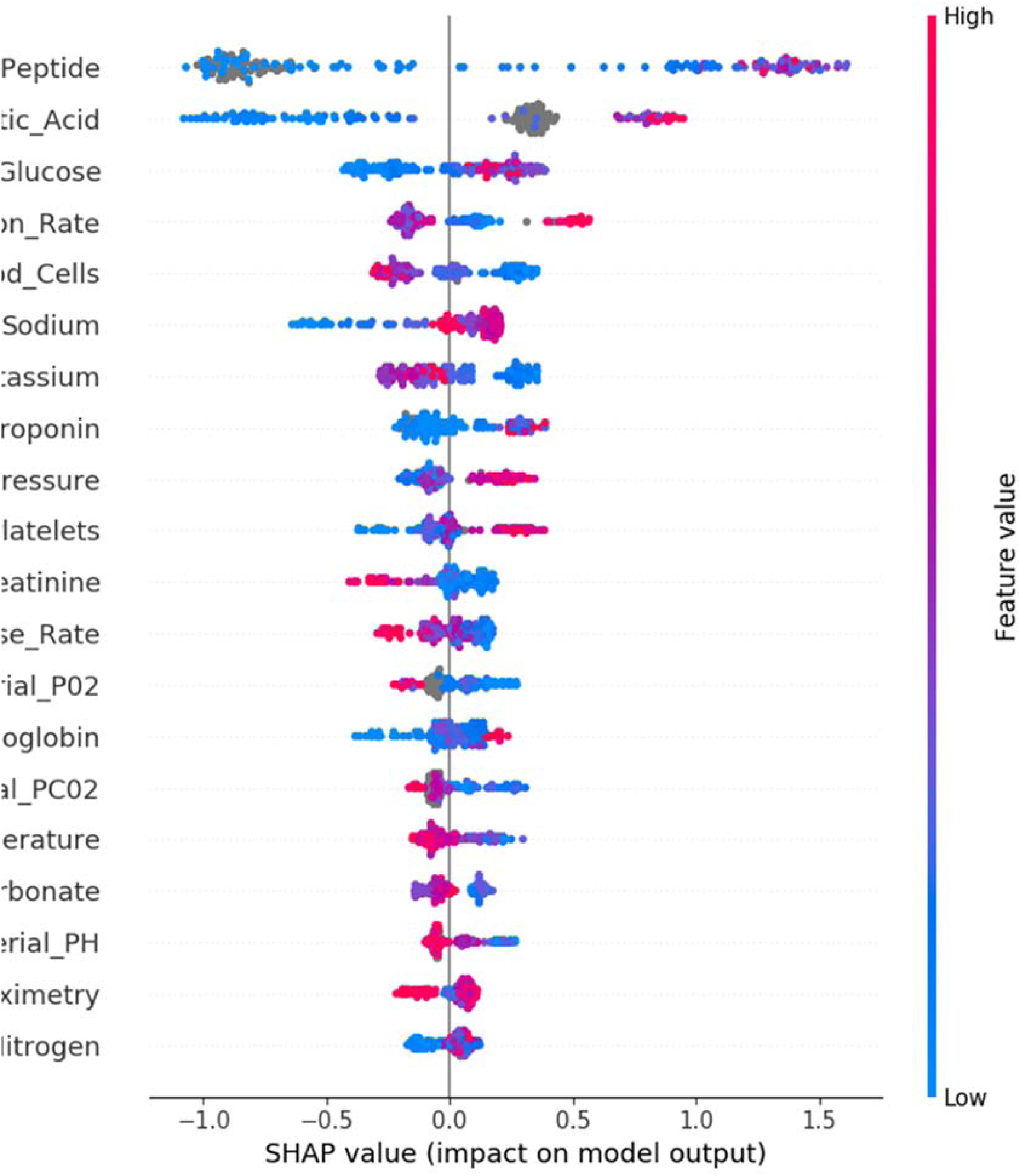
SHAP Feature importance plot for clinical features on the “cardiac” labeling task.

**Figure 7:**
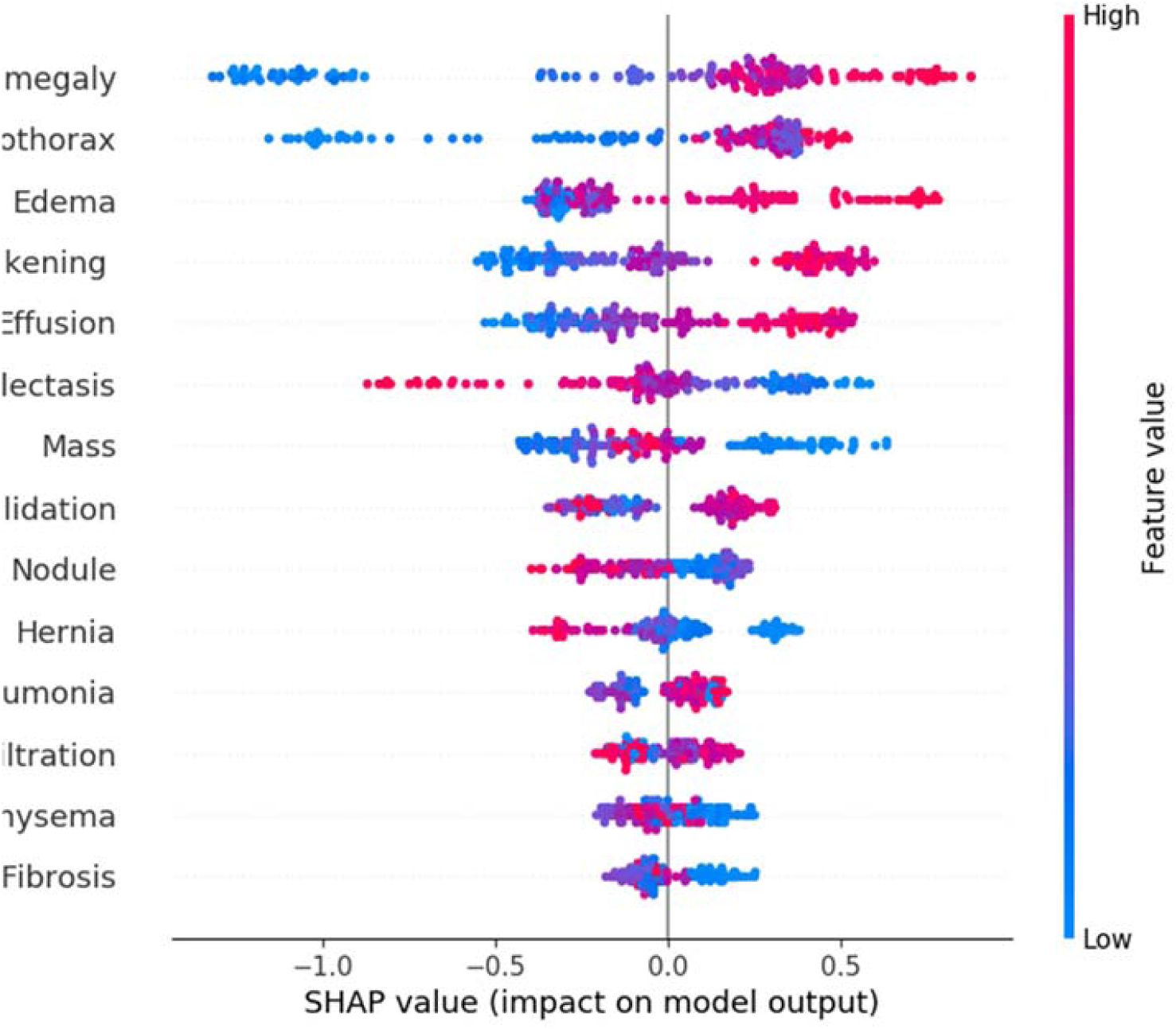
SHAP Feature importance plot for image features on the “cardiac” labeling task.

The model’s evaluation of imaging features for cardiac causes are less intuitive. The model highly values cardiomegaly, effusion, and edema as predictive of a cardiac cause. All these radiographic findings can be present in heart failure [5, 8]. However, the model’s use of the pneumothorax and pleural thickening features as predictive of heart failure, do not make clinical sense. The model may be using these features to evaluate for the presence of Kerley B lines. These lines are commonly associated with heart failure and are adjacent to the pleura [15].

#### 3.2.3 Feature Comparison

Table 3 shows the top five clinical features for each labeling task. The top features were determined by summarizing all absolute values of SHAP values by features and then ranking features based on the sum.

**Table 3:** Top 5 clinical features by labeling task.

| Rank | Infection | Cardiac |
| --- | --- | --- |
| 1 | White_Blood_Cells | Brain_Natriuretic_Peptide |
| 2 | Lactic_Acid | Lactic_Acid |
| 3 | Temperature | Glucose |
| 4 | Pulse_Rate | Respiration_Rate |
| 5 | Hemoglobin | White_Blood_Cells |

Table 4 shows the top five clinical panels for each labeling task. The top panels were determined by further summarizing and ranking the per-feature SHAP value magnitudes. A panel with more components is potentially favored in ranking as more values are added together. The single-component feature B-NP ranks 1st in the Cardiac experiment suggests that it is a very strong indicator.

**Table 4:** Top 5 clinical panels comparison. The number of features in each panel is shown in parentheses.

| Rank | Infection | Cardiac |
| --- | --- | --- |
| 1 | Vitals (6) | B-NP (1) |
| 2 | CBC (3) | BMP (7) |
| 3 | BMP (7) | Vitals (6) |
| 4 | ABG (4) | Lactic Acid (1) |
| 5 | Lactic Acid (1) | CBC (3) |

Table 5 shows the top five imaging features, labeled by their corresponding CheXNet label [14] for each labeling task. The top features were determined by SHAP analysis as previously described.

**Table 5:**
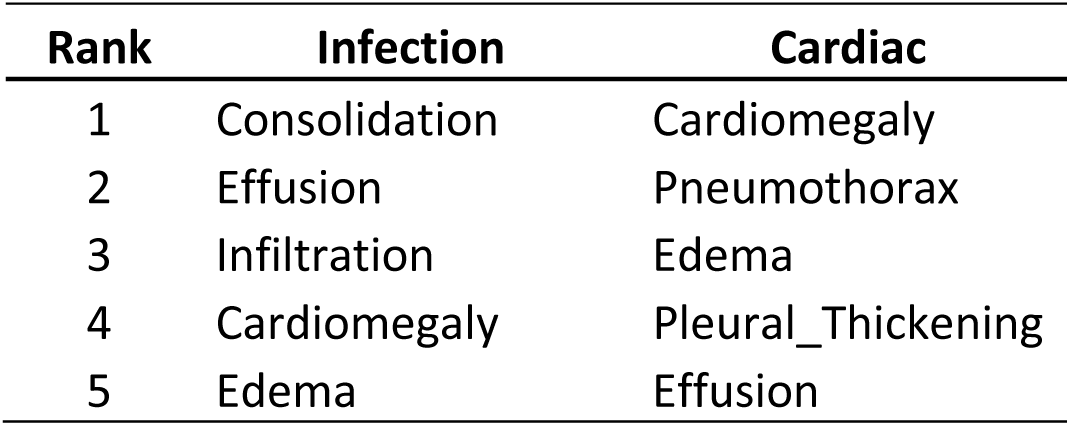
Top 5 image features comparison.

##### Feature Importance

As seen in Table 3, the model ranks lactic acid measurements as its second most important feature for both infectious and cardiac causes of respiratory distress. Lactic acidosis is usually caused by global hypoperfusion which could be secondary to cardiac (cardiogenic shock) or infectious (sepsis) causes [7]. This suggests that, in our dataset, patients with cardiac causes of acute respiratory distress are more likely to also present with lactic acidosis than those with infectious causes, or that they are likely to develop lactic acidosis sooner. This makes clinical sense as an infection in the lungs need not have systemic effects to cause respiratory distress whereas heart failure is expected to have systemic effects.

The model seems to view the classifications of infection and cardiac causes of acute respiratory distress as somewhat dichotomous. For instance, high values of lactic acid are associated with cardiac causes and low values are associated with infectious causes, and this value is ranked as the second most important for both classifications. Similarly, white blood cell count is the most important laboratory value for infection and the fifth most important for cardiac, with high values associated with infection and low values associated with cardiac.

As seen in Table 5, we observe imaging features shared between the infection and cardiac classifications, with edema, cardiomegaly, and effusion in the top 5 features for both categories. Even though effusion can sometimes be associated with complicated pneumonia [8], the model treats all of these features as favoring cardiac causes while disfavoring infectious causes, reinforcing the model’s overall dichotomous view of these disease processes.

## Summary and Discussion

### Current Performance of the Model

We show that a combination of imaging and clinical features improved overall performance of XGBoost on predicting both infectious and cardiac causes of acute respiratory distress. For the infection labeling task, the combined model performed best in 3 of the 5 cross validation folds, and performed slightly better on average. In this task, the performance was only marginally better than clinical features alone, perhaps due to the higher variance in visual presentation of infectious conditions. In the cardiac labeling task, the combined model performed best in 4 out of 5 folds. Interestingly, in this task the imaging model alone significantly underperformed the clinical model, but the image features provided a larger overall improvement when added to the clinical features than we saw on the infection labeling task. The combination seems to improve the consistency of XGBoost on prediction of cardiac causes.

### Future Performance Expectations

The main limitation on this model’s current performance is the relatively small number of example cases. The dataset of 171 patients is far below an ideal number for training. However, we are continuing to expand our dataset. As our dataset grows, we expect significant performance improvements. We are also exploring new image model formulations that make use of “localization” annotations we were able to collect on our dataset. These annotations should allow us to provide addition feedback to the image model to serve as a forcing function for an attention mechanism. With an updated model and by expanding our dataset to hundreds of cases, we expect accuracy to make significant improvements in performance.

### Future Task Expansion

This project was started before the recent SARS-CoV-2 pandemic. As we move forward with development, we will explore upgrading the model to include a SARS-CoV-2 specific classification with COVID-19 patients data. This would allow physicians to use the same software to diagnose cases of SARS-CoV-2 pneumonia. We expect our model to be able to perform this task with a high accuracy as other research teams have had success with this problem [16]. This would also support our goal of improving antibiotic stewardship among physicians as SARS-CoV-2 pneumonia does not benefit from antibiotic therapy [11].

### Future Research

In our next phase of research, we will allow our collaborating resident physicians to apply the model to new patients and help guide decision making. This will allow us to evaluate the model’s efficacy in improving patient outcomes and reducing antibiotic use.

### Web Access to Model

We have published the current version of this model on the Internet for evaluation purposes. Physicians will be able to enter the data and receive a prediction through a web interface for research purposes. Eventually, our goal is to aid emergency room clinicians in planning treatment strategies, although clinical evaluation and approval is required before it can be used as a diagnostic and planning tool. It is available at: http://nbttranslationalresearch.org/.

## Data Availability

The clinical data and image data for this study were collected and prepared by the residents and researchers of the Joint Translational Research Lab of Arkansas State University (A-State) and St. Bernards Medical Center (SBMC) Internal Medicine Residency Program. As data collection is on-going for the project stage-II of clinical testing, raw data is not currently available for data sharing to the public.

## Acknowledgment

This research work was partially supported by National Institute of Health NCI grant U01CA187013, and National Science Foundation with grant number 1452211, 1553680, and 1723529, National Institute of Health grant R01LM012601, as well as was partially supported by National Institute of Health grant from the National Institute of General Medical Sciences (P20GM103429).

## Notes

### Competing Interest Statement

The authors have declared no competing interest.

